# Plasma acetylated α-synuclein as a novel quantitative biomarker for Parkinson’s disease

**DOI:** 10.64898/2025.12.28.25342986

**Authors:** Ryoji Goto, Harutsugu Tatebe, Ayami Okuzumi, Gen Matsumoto, Kenji Tagai, Sayo Matsuura, Shunichi Niiyama, Taiji Tsunemi, Taku Hatano, Haruki Nakagawa, Taro Tachibana, Fukiko Kitani-Morii, Takashi Kasai, Fuyuki Kametani, Masato Hasegawa, Wataru Satake, Tatsushi Toda, Yuko Kataoka, Rin Yanai, Masafumi Shimojo, Hironobu Endo, Makoto Higuchi, Takahiko Tokuda

## Abstract

**Background:** Aggregation of α-synuclein is a central pathological feature of Parkinson’s disease (PD), yet reliable and broadly applicable fluid biomarkers reflecting disease-relevant α-synuclein biology remain limited. We aimed to establish acetylated α-synuclein (Ac-αSyn), the predominant proteoform *in vivo*, as a novel biomarker for PD and to evaluate its diagnostic utility based on a sensitive immunoassay.

**Methods:** Using a single molecule array technique capable of quantitatively detecting N-terminally acetylated α-synuclein, plasma Ac-αSyn levels were measured in 110 samples obtained from 52 patients with PD, 24 patients with multiple system atrophy (MSA), and 34 healthy controls (HCs). In a subset of PD patients, plasma Ac-αSyn measurements and ^123^I-metaiodobenzylguanidine (MIBG) cardiac scintigraphy were performed in the same individuals, enabling direct comparison between these two testing modalities. Ac-αSyn levels were also quantified in 91 cerebrospinal fluid (CSF) samples obtained from 51 patients with PD, 25 patients with MSA, and 15 non-parkinsonian disease controls (DCs).

**Results:** Plasma Ac-αSyn levels robustly differentiated PD from both MSA and HCs (*p* < 0.0001). Receiver operating characteristic analysis demonstrated high diagnostic performance (area under the curve [AUC] = 0.89 for PD vs MSA; AUC = 0.94 for PD vs HCs), comparable to established imaging biomarkers. In the same individuals, plasma Ac-αSyn levels correlated with the heart-to-mediastinum ratio derived from MIBG cardiac scintigraphy. CSF Ac-αSyn levels also clearly differentiated PD from both MSA and DCs (*p* < 0.0001), with high diagnostic performance (AUC = 0.85 for PD vs MSA; AUC = 0.93 for PD vs DCs), supporting the biological relevance of plasma Ac-αSyn as a biomarker.

**Conclusion:** This study identifies Ac-αSyn in plasma as a novel biomarker for PD, enabled by quantitative immunoassay-based detection. Plasma Ac-αSyn represents a practical and minimally invasive biomarker that supports biology-based diagnosis of PD and discrimination from MSA.

## 1. Background

The accumulation of α-synuclein is a pathological hallmark of several neurodegenerative disorders, including Parkinson’s disease (PD), dementia with Lewy bodies (DLB), and multiple system atrophy (MSA)^1^. Abnormal α-synuclein constitutes the core component of Lewy bodies and Lewy neurites in PD and DLB ^2–4^, whereas it is the principal constituent of glial cytoplasmic inclusions in MSA^5^. The aggregation of α-synuclein is considered to contribute to neuronal loss in these α-synucleinopathies^6^, positioning α-synuclein as a central pathological marker and as a major target for disease-modifying therapies^7,8^.

In parallel with these pathological insights, a biological definition of PD has been proposed recently^9,10^. This framework reflects the limited specificity of clinical symptoms and the recognition that PD evolves along a continuous spectrum, including a prolonged preclinical phase preceding overt motor manifestations. A biology-based diagnosis grounded in disease-defining molecular pathology is therefore expected to enable earlier detection, improve diagnostic precision, and accelerate therapeutic development by aligning interventions with underlying disease mechanisms.

Despite the central role of α-synuclein in PD pathogenesis, quantitative assessment of α-synuclein pathologies in living patients remains challenging. PD-associated neurodegeneration and dopaminergic dysfunction can be evaluated using established imaging modalities such as dopaminergic SPECT or PET; however, technologies that directly capture α-synuclein abnormalities are limited. Recent advances in imaging have led to the development of α-synuclein PET tracers, including ^18^F-C05-05, which can visualize α-synuclein deposits in the midbrain of patients with PD and DLB^11^. Nevertheless, the utility of these tracers for early diagnosis and longitudinal disease monitoring remains to be established.

Fluid biomarkers have emerged as a complementary approach to imaging. In particular, seed amplification assays (SAAs) have demonstrated high diagnostic sensitivity for detecting misfolded α-synuclein seeds in cerebrospinal fluid (CSF)^10^ and, more recently, in serum samples from patients with PD and MSA^12^. While these assays provide remarkable sensitivity, their clinical implementation is constrained by limited scalability, dependence on specialized equipment, inter-laboratory variation, lack of standardized reference materials, and their fundamentally qualitative nature. These limitations underscore the need for quantitative, scalable, and widely applicable blood-based biomarkers that directly reflect α-synuclein pathobiology.

Despite substantial attempts to quantify α-synuclein in blood using immunoassays, these efforts have yielded inconsistent results, including conflicting reports on whether α-synuclein levels are elevated in α-synucleinopathies compared with healthy controls^13–16^. Although variability in patient characteristics and sample handling, especially the influence of hemolysis, has been suggested as a contributing factor, inconsistencies persist even after controlling for these variables^17,18^, highlighting intrinsic limitations of existing immunoassay strategies. Assays targeting oligomeric or phosphorylated α-synuclein have also been expolored^19,20^, but their diagnostic performance has remained insufficient, particularly for distinguishing PD from MSA.

α-Synuclein undergoes multiple post-translational modifications (PTMs)^21^, among which acetylation of the N-terminal initiator Met residue (N-terminal acetylation) is of particular biological relevance. N-terminal acetylation occurs in the majority of eukaryotic proteins and is considered irreversible. In the case of α-synuclein, this modification affects its interactions with membranes and the equilibrium between the monomeric and tetrameric states^22^ and is considered to represent the functional form of the protein^23^, priming other PTMs^23^. Nuclear magnetic resonance and electron paramagnetic resonance spectroscopy revealed that N-terminally acetylated α-synuclein is the predominant native state *in vivo*^24^. Mass spectrometry (MS) has consistently detected this modification in postmortem brain tissues from healthy controls and patients with Lewy body disease^25^ and MSA^26,27^, and only the N-terminal acetylated form was present in brain samples^28,29^. Moreover, MS studies documented N-terminal acetylated α-synuclein as the most abundant proteoform in blood samples of healthy controls and patients with PD^30,31^. Despite its biological predominance and relevance, N-terminally acetylated α-synuclein has remained largely underexplored as a fluid biomarker^32^. In the present study, we established acetylated α-synuclein as a novel biomarker for PD. Using a quantitative immunoassay platform, we demonstrate that acetylated α-synuclein can be reliably measured in plasma and CSF and enables discrimination of PD from MSA and controls, supporting its utility for biology-based diagnosis of PD.

## 2. Methods

### Subject characterization and sample collection

To evaluate acetylated α-synuclein as a diagnostic biomarker, its levels were measured with a novel in-house assay in human plasma and CSF samples. Two independent sample sets were analyzed: a plasma cohort and a CSF cohort, with paired plasma samples available for a subset of participants.

The plasma cohort comprised samples from patients with PD (n = 52) and MSA [n = 24; 23 MSA with predominant parkinsonism (MSA-P) and 1 MSA with predominant cerebellar ataxia (MSA-C)], collected at Juntendo University Department of Neurology and Neurosurgery. Plasma samples from healthy controls (HCs; n = 35) without neurodegenerative disorders were collected at the National Institutes for Quantum Science and Technology (QST).

The CSF cohort included paired CSF and plasma samples from patients with PD and comorbid idiopathic normal pressure hydrocephalus (iNPH; n = 51) and from patients with MSA (n = 6; 5 MSA-P and 1 MSA-C), collected at Juntendo University Department of Neurology and Neurosurgery. Additional CSF samples were obtained from patients with MSA (n = 19; 3 MSA-P and 16 MSA-C) and from disease controls (DCs; n = 15) without neurodegenerative disorders who underwent lumbar puncture as part of the diagnostic evaluation at the Department of Neurology, Graduate School of Medical Science, Kyoto Prefectural University of Medicine. The diagnostic criteria for PD, MSA, and iNPH were based on the Movement Disorder Society (MDS)-sponsored PD clinical criteria^33^, Gilman’s criteria^34^, and the established clinical guidelines for iNPH^35^, respectively.

Plasma samples were collected in ethylenediaminetetraacetic acid (EDTA)-containing tubes and stored in polypropylene vials at – 80[°C. CSF samples were collected in polypropylene tubes, centrifuged at 3,000×g for 15 minutes at 4°C to remove cellular debris, and stored at –80[°C. All samples were subsequently analyzed at QST.

### Ethics, consent and permissions

The study was approved by the Ethics Committee of all institutions. Written informed consent was obtained from all participants before enrollment. The study procedures were performed in accordance with the Declaration of Helsinki.

### Immunoassay protocols

After excluding plasma and CSF samples with evidence of hemolysis, acetylated α-synuclein concentrations were measured using a novel ultrasensitive immunoassay on the Simoa HD-X analyzer (Quanterix) at the Advanced Neuroimaging Center, QST. The assay was designed to selectively detect N-terminally acetylated human α-synuclein.

A monoclonal antibody recognizing the N-terminal acetylated region of human α-synuclein was conjugated to paramagnetic beads (#103207; Quanterix) and used as the capture antibody. A commercially available mouse monoclonal antibody (Syn-1; #610787; BD Bioscience) was employed as the detector antibody after biotinylation according to the manufacturer’s instructions. A custom synthetic peptide containing the N-terminal acetylation site and mid-portion epitope of α-synuclein (molecular weight 3313.3 Da; Eurofins Genomics) was used as the assay calibrator.

Standard curves were generated in duplicate using serial dilutions of the custom peptide in calibration buffer composed of 85.5% Homebrew sample diluent (#101351; Quanterix) supplemented with 0.9% bovine serum albumin (#60-00020-10; pluriSelect Life Science) and 0.9% protease inhibitor (#539131-10VLCN; Sigma-Aldrich). All plasma samples stored at −80[°C were thawed, vortexed, and diluted 1:4 with sample buffer containing 93.6% Homebrew sample diluent (#101351; Quanterix) supplemented with 1% bovine serum albumin (#60-00020-10; pluriSelect Life Science), 1% protease inhibitor (#539131-10VLCN; Sigma-Aldrich).

For the assay, 40 μL of either sample or calibrator was incubated with the antibody-coated beads for 30 minutes. After washing the beads, the biotinylated detector antibody was added and incubated, followed by additional bead washes. Subsequently, 100 μL of streptavidin-conjugated β-galactosidase (SBG, #100439; Quanterix) was added and incubated. After another bead wash, resorufin β-D-galactopyranoside (RGP; #103159; Quanterix) was applied, and the resuspended beads were transferred to a multiwell disc array for imaging. Fluorescent signals were quantified and converted to average enzyme per bead (AEB) values as previously described^36^, and acetylated α-synuclein concentrations were calculated by four-parameter logistic regression using the calibrator standards.

Detailed procedures for antibody preparation, affinity verification, and analytical validation of the acetylated α-synuclein assay are provided in the eMethods of the Supplement.

### Hemolysis evaluation

Hemolysis was assessed by visual inspection and spectrophotometric measurement of the hemolysis coefficient at 414 nm (UV_–_ Vis module) using a NanoDrop-1000 spectrophotometer (Thermo Fisher Scientific). Measurements were performed in duplicate using 1.5 μL of plasma or CSF aspirated from the middle layer of the sample tube, as described elsewhere^37^. Samples with a hemolysis coefficient exceeding an established threshold were excluded from subsequent immunoassay analyses.

### Radiological assessment

^123^I-metaiodobenzylguanidine (MIBG) cardiac scintigraphy and dopamine transporter (DAT) single photon emission computed tomography (SPECT) images were acquired and evaluated as previously described^38,39^.

### Statistical analysis

Demographic and clinical characteristics were summarized using nonparametric statistics. Group differences in continuous variables were assessed using the Mann–Whitney U test, and categorical variables were compared using the chi-squared test. Differences in plasma acetylated α-synuclein levels among diagnostic groups were evaluated using one-way analysis of variance (ANOVA). Pearson’s correlation test was used to examine correlations between plasma acetylated α-synuclein levels and age or disease duration, whereas Spearman’s rho test was used to examine correlations with clinical scores. Diagnostic performance was evaluated by receiver operating characteristic (ROC) analysis, with the area under the curve (AUC) and its standard error estimated using a nonparametric method^40^. Optimal cutoff values were determined using Youden’s index, i.e., J = max(sensitivity + specificity–1)^41^. Comparisons between AUCs were performed using the DeLong method.

For plasma samples with acetylated α-synuclein levels below the limit of detection (LOD), values were imputed to the LOD for statistical analysis. All statistical analyses were performed using Prism software (version 10.4.1; GraphPad Software, San Diego, CA, USA), and statistical significance was defined as a two-sided p value < 0.05.

## 3. Results

### Subjects and sample characteristics

In the plasma cohort comprising patients with PD, MSA, and HCs, all samples except for one HC sample showed hemolysis coefficients below the predefined exclusion threshold and were included in the immunoassays. In the CSF cohort comprising patients with PD, MSA, and DCs, along with paired plasma samples from all PD patients and a subset of MSA patients, all CSF samples and all paired plasma samples from patients with MSA showed no evidence of hemolysis. In contrast, 35 plasma samples from patients with PD exceeded the hemolysis threshold and were removed from subsequent measurements, resulting in 16 evaluable paired plasma samples from this group.

Demographic and clinical characteristics of the analyzed samples are summarized in Table 1.

**Table 1.** Demographic data of the participants.

| **A: Participants for plasma measurement** | Parkinson's disease | Multiple system atrophy | Healthy control |
| --- | --- | --- | --- |
| Total number of plasma samples | 52 | 24 | 35 |
| Number of non-hemolyzed plasma samples used for immunoassay | 52 | 24 | 34 |
| Age | 62.8 ± 9.0 | 61.0 ± 9.0 | 52.1 ± 8.6 |
| Gender (male/female) | 34/18 | 13/11 | 12/22 |
| Disease duration (years) | 9 (1–31) | 4 (1–16) | N/A |
| MDS-UPDRS part 3 | 19 (2–58) | 52.5 (11–108) | N/A |
| Yahr | 2 (1–4) | 3 (2–5) | N/A |
| MMSE | 29 (14–30) | 28 (20–30) | N/A |
| MoCA-J | 25 (9–30) | N/A | N/A |
| FAB | 15 (4–18) | N/A | N/A |
| Open Essence | 4 (0–10) <sup>[1]</sup> | N/A | N/A |
| MIBG cardiac scintigraphy |  |  |  |
| early acquisition | 1.7 (1.1–3.8) <sup>[2]</sup> | 3.0 (2.4–3.9) <sup>[3]</sup> | N/A |
| late acquisition | 1.4 (1.0–3.7) <sup>[2]</sup> | 3.0 (2.2–4.3) <sup>[3]</sup> | N/A |
| washout rate | 15.7%<br>(–9.8% – 29.2%) <sup>[2]</sup> | –4.8%<br>(–32.7% – 28.7%) <sup>[3]</sup> | N/A |
| DAT SPECT SBR | 2.9 ± 1.4 | 2.8 ± 1.7 <sup>[4]</sup> | N/A |
| **B: Participants for the** | Parkinson's disease | Multiple system | Disease control |

| measurement of CSF and paired plasma |  | atrophy |  |
| --- | --- | --- | --- |
| Number of CSF samples | 51 | 25 | 15 |
| Total number of plasma samples | 51 | 6 | 0 |
| Number of non-hemolyzed paired plasma samples used for immunoassay | 16 | 6 | 0 |
| Age | 75.5 ± 6.8 | 63.1 ± 7.8 | 50.9 ± 16.9 |
| Gender (male/female) | 31/20 | 13/12 | 7/8 |
| Disease duration (years) | 7.0 (1.0–31) | 2.8 (0.9–6.2) | N/A |
| MDS-UPDRS part 3 | 33 (3–79) | 32 (13–100) <sup>[5]</sup> | N/A |
| Yahr | 3 (1–5) | 3 (2–5) | N/A |
| ICARS | N/A | 24.5 (16–49) <sup>[5]</sup> | N/A |
| MMSE | N/A | 28 (20–30) <sup>[6]</sup> | N/A |

Values are expressed as means ± SD or medians (ranges). The items were analyzed in [1] 51, [2] 46, [3] 21, [4] 23, [5] 10, and [6] 19 patients.
MDS-UPDRS, Movement Disorder Society-Unified Parkinson's Disease Rating Scale; MMSE, Mini-Mental State Examination; MoCA-J, Japanese version of Montreal Cognitive Assessment; FAB, Frontal Assessment Battery; MIBG, <sup>123</sup>I- metaiodobenzylguanidine; DAT SPECT, dopamine transporter single-photon emission computed tomography; SBR, specific binding ratio; ICARS, International Cooperative Ataxia Rating Scale; N/A, not applicable.

### Quantification of acetylated α-synuclein in plasma obtained from patients with PD and MSA and HCs

Assay characteristics, including antibody reactivity to acetylated α-synuclein, standard curve performance, and analytical validation, are described in the eResults section, eTables 1-4, and eFigures 1-4 of the Supplement. Using this assay, plasma acetylated α-synuclein levels were significantly higher in patients with PD than in patients with MSA (*p* < 0.0001) and HCs (*p* < 0.0001), whereas no significant difference was observed between the MSA and HC groups (*p* = 0.41) (Fig. 1a).

**Figure 1.**
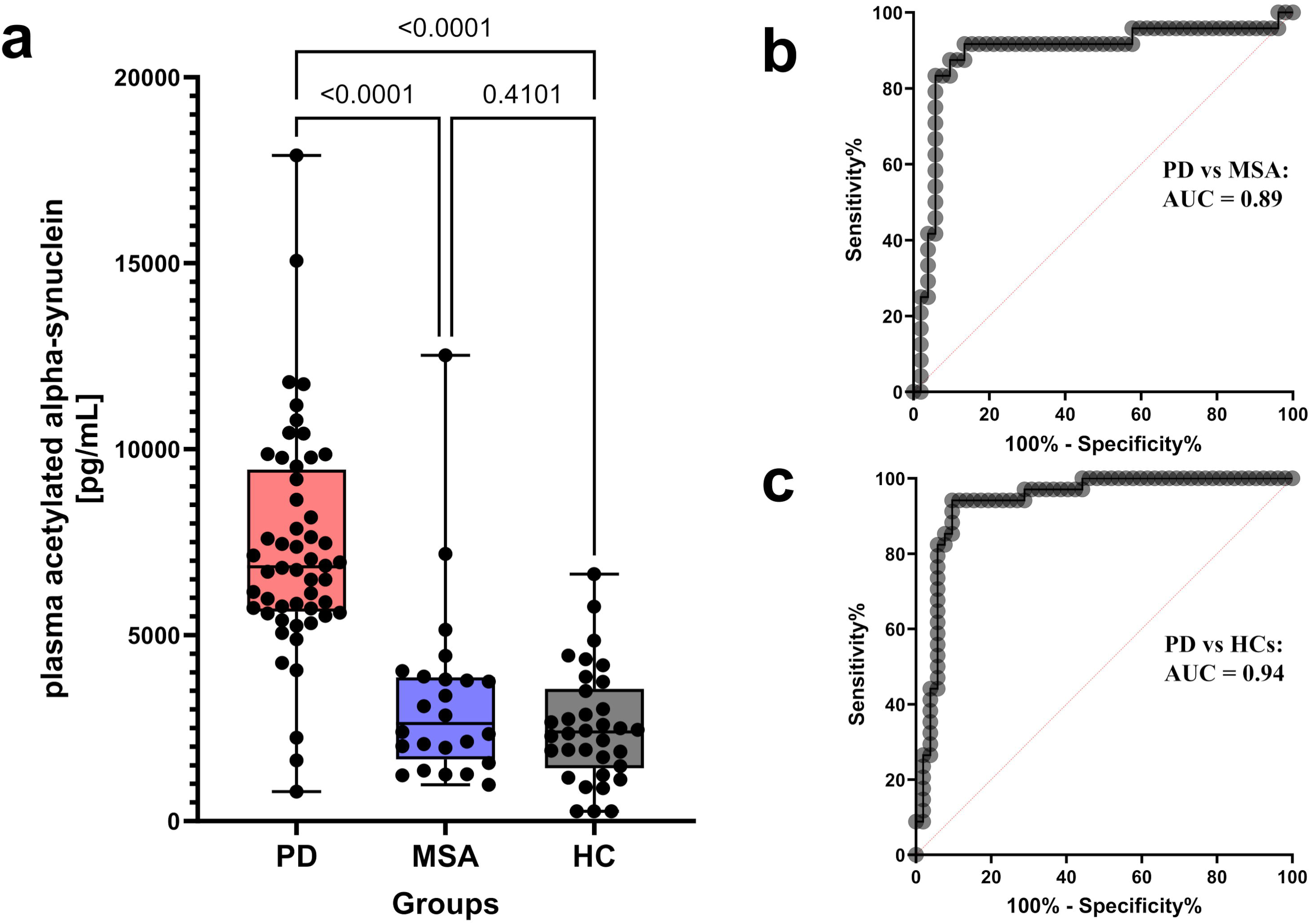
The diagnostic performance of plasma acetylated α-synuclein with our assay. **(a)** Box-and-whisker plot showing the concentrations of plasma acetylated α-synuclein in patients with Parkinson’s disease (PD) (n = 52), in those with multiple system atrophy (MSA) (n = 24), and in healthy controls (HCs) (n = 34). Plasma acetylated α-synuclein levels were significantly higher in those with PD compared with those with MSA (*p* < 0.0001) and HCs (*p* < 0.0001). **(b)** Receiver operating characteristic (ROC) curve of plasma acetylated α-synuclein levels for distinguishing patients with PD from MSA (area under the curve (AUC) = 0.89; sensitivity = 87.5%, specificity = 90.4%, cutoff value = 4,668 pg/mL). **(c)** ROC curve of plasma acetylated α-synuclein levels for distinguishing patients with PD from HCs (AUC = 0.94; sensitivity = 94.1%, specificity = 90.4%, cutoff value = 4,872 pg/mL).

**Figure 2.**
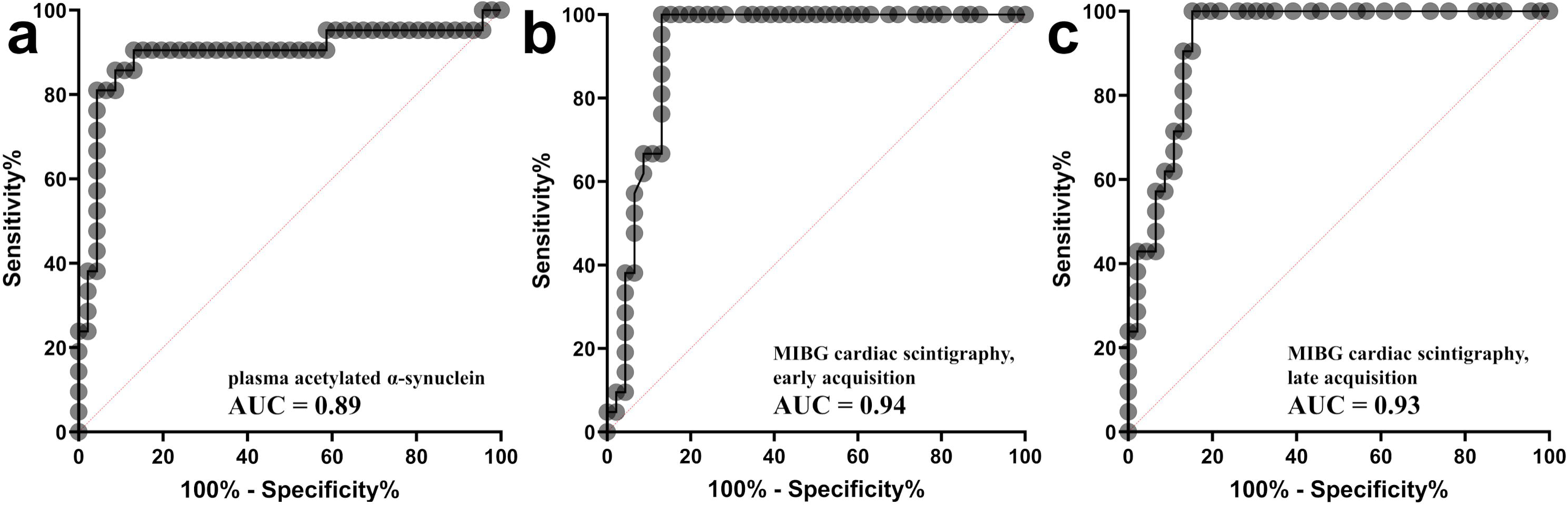
ROC curves for distinguishing PD from MSA among subjects who underwent. ^123^**I-metaiodobenzylguanidine (MIBG) cardiac scintigraphy. (a)** ROC curve of plasma acetylated α-synuclein levels. The area under the curve (AUC) was 0.89 (95% confidence interval (CI): 0.79–1.00). **(b)** ROC curve of the heart-to-mediastinum (H/M) ratio of early acquisition of MIBG cardiac scintigraphy. The AUC was 0.94 (95% CI: 0.86–0.99). **(c)** ROC curve of the heart-to-mediastinum (H/M) ratio of late acquisition of MIBG cardiac scintigraphy. The AUC was 0.93 (95% CI: 0.88–0.99).

**Figure 3.**
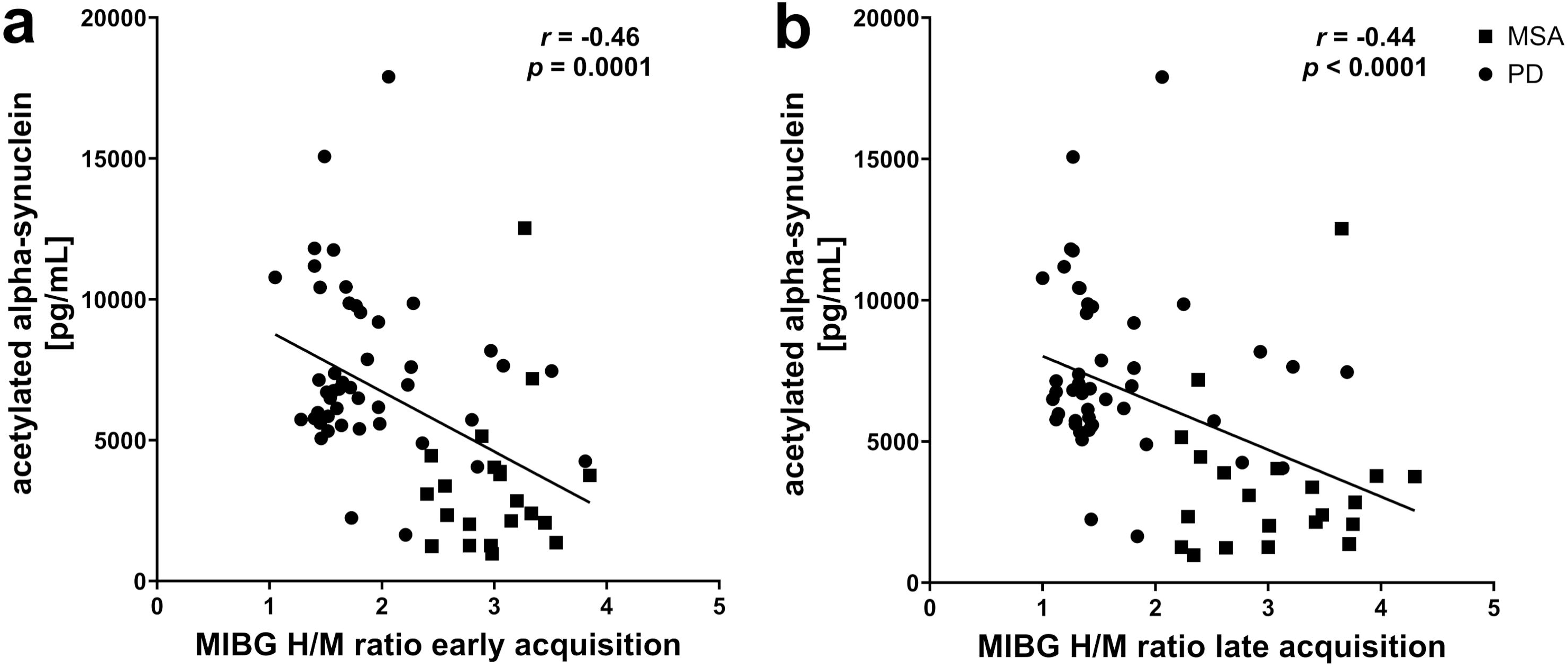
Correlation between plasma acetylated α-synuclein levels and the H/M ratio of MIBG cardiac scintigraphy. In the analysis of patients with α-synucleinopathy, plasma acetylated α-synuclein levels correlated with the H/M ratio of MIBG cardiac scintigraphy in early acquisition (*r* = –0.46, *p* < 0.0001; panel **a**) and late acquisition (*r* = –0.44, *p* < 0.0001; panel **b**).

**Figure 4.**
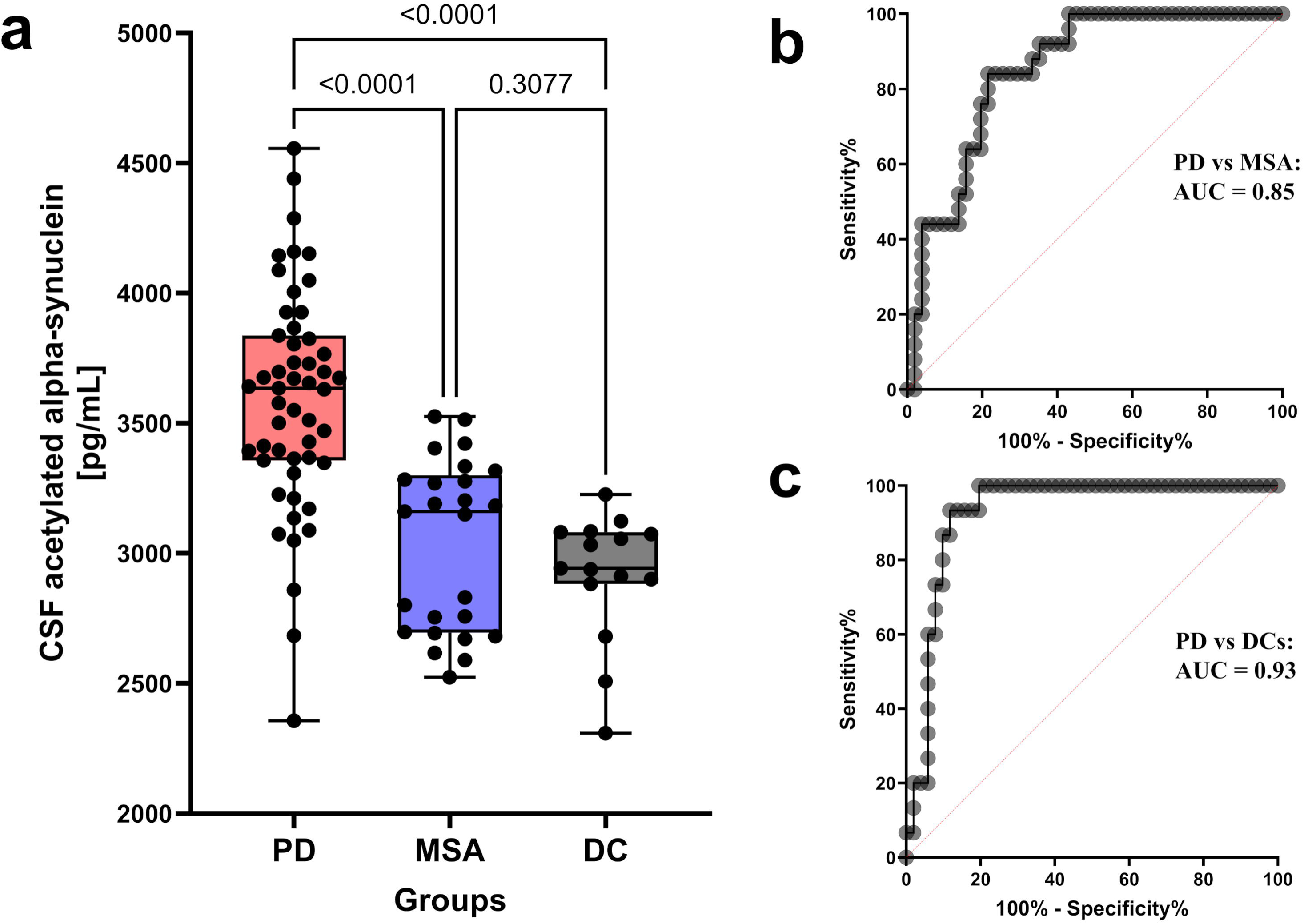
The diagnostic performance of CSF acetylated α-synuclein with our assay. **(a)** Box-and-whisker plot showing the concentrations of CSF acetylated α-synuclein in patients with Parkinson’s disease (PD) (n = 51), those with multiple system atrophy (MSA) (n = 25), and disease controls (DCs) (n = 15). CSF acetylated α-synuclein levels were significantly higher in those with PD compared with those with MSA (*p* < 0.0001) and DCs (*p* < 0.0001). **(b)** Receiver operating characteristic (ROC) curve of CSF acetylated α-synuclein levels for distinguishing patients with PD from MSA (AUC = 0.85; sensitivity = 84.0%, specificity = 78.4%, cutoff value = 3,341 pg/mL). **(c)** ROC curves of CSF acetylated α-synuclein levels for distinguishing patients with PD from DCs (AUC = 0.93; sensitivity = 93.3%, specificity = 88.2%, cutoff value = 3,129 pg/mL).

ROC analysis demonstrated high diagnostic performance of plasma acetylated α-synuclein for distinguishing PD from MSA (AUC = 0.89; cutoff value = 4,668 pg/mL; Fig. 1b) and from HCs (AUC = 0.94; cutoff value = 4,872 pg/mL; Fig. 1c).

To benchmark the diagnostic performance against an established clinical biomarker, plasma acetylated α-synuclein was compared with MIBG cardiac scintigraphy in individuals who underwent both assessments (46 patients with PD and 21 patients with MSA). In this subset, plasma acetylated α-synuclein discriminated PD from MSA with an AUC of 0.89 (cutoff value = 4,668 pg/mL; Fig. 2a), while MIBG scintigraphy achieved AUCs of 0.94 and 0.93 for early and late heart-to-mediastinum (H/M) ratio acquisitions, respectively (cutoff value = 2.2 for both acquisitions; Fig. 2b, c). There was no significant difference in diagnostic performance between plasma acetylated α-synuclein and MIBG scintigraphy, as assessed by the DeLong test (*p* = 0.66 for early acquisition and *p* = 0.53 for late acquisition). Consistent with this finding, plasma acetylated α-synuclein levels inversely correlated with the H/M ratio of MIBG cardiac scintigraphy in patients with PD and MSA (*r* = –0.46, *p* = 0.0001 for early acquisition; *r* = –0.44, *p* < 0.0001 for late acquisition; Fig. 3), indicating an association between plasma acetylated α-synuclein levels and cardiac sympathetic denervation.

In contrast, no significant correlation was observed between plasma acetylated α-synuclein levels and specific binding ratio of DAT SPECT, either in the analysis including patients with PD and MSA or in the analysis restricted to those with PD. Plasma acetylated α-synuclein levels also did not show significant correlations with clinical severity measures in patients with PD (eTable 5).

### Quantification of acetylated α-synuclein in CSF and paired plasma

CSF acetylated α-synuclein levels were significantly higher in patients with PD than in patients with MSA and DCs (*p* < 0.0001 for both comparisons; Fig 4a). Although mean CSF acetylated α-synuclein levels were numerically higher in the MSA group than in the DC group, the difference was not statistically significant (*p* = 0.28).

ROC analysis demonstrated that CSF acetylated α-synuclein discriminated PD from MSA with an AUC of 0.85 (cutoff value = 3,341 pg/mL; Fig. 4b) and from DCs with an AUC of 0.93 (cutoff value = 3,129 pg/mL; Fig. 4c).

Correlation analyses revealed no significant association between CSF acetylated α-synuclein levels and clinical measures in patients with PD (eTable 6) or in DCs, including age (*r* = –0.16, *p* = 0.56). In patients with MSA, CSF acetylated α-synuclein levels did not differ between samples collected from Juntendo University and Kyoto Prefectural University of Medicine (*p* = 0.14), nor between MSA-P and MSA-C subtypes (*p* = 0.3440; eFigure 5). No significant correlation with clinical variables was observed in the MSA group (eTable 7), with the exception of a negative correlation with Mini-Mental State Examination scores (*r* = –0.601, *p* = 0.0065; eFigure 6).

We also investigated the correlation between CSF and plasma acetylated α-synuclein levels in the same patients. No significant correlations were observed between CSF and non-hemolyzed plasma acetylated α-synuclein levels in patients with PD (*r* = –0.12, *p* = 0.65) or in those with MSA (*r* = – 0.10, *p* = 0.86) (eFigure 7).

## 4. Discussion

In this study, we established acetylated α-synuclein as a novel fluid biomarker for PD by demonstrating its robust and quantitative detectability in both plasma and CSF, and its ability to reliably discriminate PD from MSA and control groups. Plasma acetylated α-synuclein levels were significantly elevated in PD compared with MSA and HCs, with diagnostic performance comparable to that of MIBG cardiac scintigraphy, a widely used clinical biomarker. CSF acetylated α-synuclein levels similarly distinguished PD from MSA and DCs, supporting the biological relevance of this proteoform across central and peripheral compartments.

The identification of acetylated α-synuclein as a blood-based biomarker has important implications in the context of the recently proposed biological definition of PD. Current clinical diagnostic frameworks^33^ incorporate neuroimaging of dopaminergic dysfunction and cardiac sympathetic denervation, which, although informative, are invasive, costly, and not readily repeatable. In contrast, our blood-based assay for plasma acetylated α-synuclein provides a minimally invasive and scalable biomarker that directly reflects disease-defining molecular pathology, thereby supporting a biology-based diagnosis of PD^9,10^ and offering a practical complement to existing imaging modalities.

Previous immunoassays targeting α-synuclein in human body fluids have yielded inconsistent results and limited diagnostic performance. Rather than representing a purely technical advance, the present findings suggest that the key determinant of diagnostic utility lies in the biological relevance of the α-synuclein species being measured. In this context, acetylated α-synuclein appears to represent a proteoform that more faithfully reflects *in vivo* α-synuclein biology.

One factor contributing to this relevance is the ability of our assay to capture α-synuclein species that are truncated at the C terminus, since the epitope of our detector antibody, Syn-1, is localized within amino acid residues 91–99^42^ and does not target the distal C-terminus. C-terminally truncated α-synuclein aggregates more readily and constitutes a substantial fraction of α-synuclein species within Lewy bodies in PD^25,28,29,43^, as well as within glial cytoplasmic inclusions in MSA^27,43^. Such truncated species have also been detected in peripheral compartments, including erythrocytes, plasma extracellular vesicles^44^, and serum^45^. Antibodies used in previous studies often target the distal C-terminus and may therefore fail to detect these abundantly present truncated isoforms^46^, particularly in the presence of PTMs or metal-dependent conformational changes that alter epitope accessibility in the C-terminal region^21^.

Equally important is the focus on N-terminally acetylated α-synuclein. N-terminal acetylation occurs co-translationally in eukaryotic cells^23,24^, and has also been suggested to represent the predominant native state of α-synuclein^24^ in the human brain^25–29^ and blood^30,31^. This modification neutralizes the positive charge at the N[terminus, promotes α-helical structure formation, and affects membrane interactions and conformational stability. In contrast, recombinant α-synuclein produced in prokaryotic systems lacks this modification and does not fully recapitulate the native α-helical conformation of the N-terminal region of human α-synuclein. Biomarker strategies that rely on recombinant standards or fail to distinguish acetylated from non-acetylated species may therefore underestimate or mischaracterize physiologically relevant α-synuclein pools. By targeting N-terminal acetylation while detecting C-terminal truncation, the present α-synuclein measurement likely captures the dominant *in vivo* proteoform across central and peripheral compartments.

Elevated acetylated α-synuclein levels in both plasma and CSF in PD further support its biological and pathophysiological relevance. Prion-like spreading of α-synuclein has been considered to lead to the development and progression of PD^47^, traditionally envisioned as propagating along synaptically connected neural networks^48,49^. However, α-synuclein pathology in PD is extensively distributed throughout peripheral organs, including the heart, kidney, and skin, and this distribution cannot be fully explained by neural connectivity alone^50^. Accumulating experimental evidence indicates that α-synuclein can cross the blood-brain barrier (BBB)^51^, and that blood-borne α-synuclein seeds can provoke aggregation within the central nervous system (CNS) after systemic administration in animal models^52,53^. Additional pathways, including renal nerve–mediated transport of blood-derived α-synuclein to the CNS, have also been proposed^54^.

In this context, the present observation that plasma levels of α-synuclein were approximately twice as high as those in CSF, together with the absence of correlation between CSF and plasma levels, supports the concept that peripheral and central α-synuclein pools are at least partially independent. These findings are consistent with a model in which blood-mediated propagation contributes to disease biology in PD, with plasma acetylated α-synuclein reflecting a compartment distinct from CSF yet biologically linked to disease propagation. Potential sources of plasma α-synuclein include transfer from the CSF, release via brain-derived extracellular vesicles^55^, and peripheral production in organs such as the gastrointestinal tract^56^ and erythrocytes^57^.

The lack of correlation between clinical measures and plasma or CSF α-synuclein levels in PD is also biologically plausible. Neuropathological studies have demonstrated that the burden of α-synuclein aggregation does not necessarily parallel the extent of neuronal loss^58,59^. It is therefore conceivable that acetylated α-synuclein levels rise early in the disease course, preceding overt neurodegeneration and symptom progression. From this perspective, elevated acetylated α-synuclein levels may represent an early pathogenic indicator rather than a marker of disease stage, underscoring their potential utility for early diagnosis.

In contrast to PD, both plasma and CSF levels of α-synuclein in MSA were comparable to those in controls. This finding, consistent with a previous report^60^, suggests a fundamental difference in the mechanisms underlying α-synuclein propagation between these disorders. In PD, increased extracellular α-synuclein and its dissemination through the blood compartment may play a prominent role. In MSA, by contrast, disease progression may be driven by enhanced aggregation propensity and increased oligodendroglial uptake of α-synuclein^61^, rather than by elevated extracellular α-synuclein concentrations. Differences in filament fold structure^62,63^, cellular vulnerability, and autonomic involvement^64^ between PD and MSA further support this distinction. The rapid spreading of α-synuclein in MSA compared with PD^12^, despite its lower concentration in body fluids, may be due to differences in the pathogenic mechanisms between PD and MSA, possibly caused by distinct filament folds^62,63^. The CNS and preganglionic autonomic nerve fibers are primarily affected in MSA, while the peripheral components of the autonomic nervous system are relatively less involved compared with Lewy body disorders^64^. Therefore, propagation of α-synuclein through the blood would be limited in MSA, and it might instead spread mainly through neuronal networks within the brain^65^. Genetic evidence is also consistent with this view, as variants that increase α-synuclein expression are strongly associated with PD phenotypes but not with MSA^66–68^.

Several limitations should be acknowledged. The CSF samples were obtained from patients with PD and comorbid iNPH. Since iNPH itself may influence acetylated α-synuclein levels in the CSF, validation in CSF samples from PD patients without such comorbid conditions is required. In addition, diagnoses in the present study were based on established clinical criteria supported by imaging findings in the absence of neuropathological confirmation. Although this approach is widely accepted in biomarker validation studies of neurodegenerative diseases^69^, pathological evaluation remains the gold standard. Validation in cohorts with pathologically confirmed diagnoses is accordingly warranted. Furthermore, the sample size was limited, and larger, multistage cohorts will be necessary to further define diagnostic thresholds and assess generalizability. Longitudinal studies will also be essential to determine whether acetylated α-synuclein can track disease progression or therapeutic responses. Further investigations should extend this approach to DLB and to prodromal populations, including individuals with REM sleep behavior disorder without motor symptoms, to evaluate its potential role in risk prediction and early intervention.

## 5. Conclusions

We establish acetylated α-synuclein as a novel fluid biomarker for Parkinson’s disease, enabling quantitative assessment in plasma and cerebrospinal fluid and reliable discrimination of PD from MSA and control subjects. By overcoming long-standing limitations in detecting disease-relevant α-synuclein species in body fluids, this biomarker provides a practical and minimally invasive approach that supports biology-based diagnosis of PD and advances the clinical translation of α-synucleinopathy biomarkers.

## Data Availability

The data supporting the conclusions of this study are presented in the main text and the Supplementary Materials. Additional data are available from the corresponding author upon reasonable request.

## Statements and Declarations

### Ethics approval and consent to participate

This study was approved by the Institutional Review Board of the National Institutes for Quantum Science and Technology and was performed in accordance with the ethical standards of the 1964 Declaration of Helsinki and its later amendments. Written informed consent was obtained from all participants or their caregivers.

### Consent for publication

All data are entirely unidentifiable and there are no details on the individuals reported.

### Competing Interests

Harutsugu Tatebe and Takahiko Tokuda are inventors of a patent application related to the immunoassay documented in this report (PCT/JP2025/35045).

### Funding

This work was supported by Japan Agency for Medical Research and Development (AMED) under Grant Number JP21dk0207055, JP24wm0625505, and JP24zf0127012, and by Japan Society for the Promotion of Science (JSPS) KAKENHI Grant Number JP25KJ0899.

### Author contributions

Ryoji Goto, Harutsugu Tatebe, Kenji Tagai, Makoto Higuchi, and Takahiko Tokuda conceived the study. Ryoji Goto and Harutsugu Tatebe developed and validated the assay with support from Sayo Matsuura, Haruki Nakagawa, Taro Tachibana, Fuyuki Kametani, Masato Hasegawa, Rin Yanai, Masafumi Shimojo, and Takahiko Tokuda. Ryoji Goto and Kenji Tagai performed the statistical analysis. Ayami Okuzumi, Shunichi Niiyama, Taiji Tsunemi, Taku Hatano, Fukiko Kitani-Morii, Takashi Kasai, Kenji Tagai, Makoto Higuchi, and Takahiko Tokuda recruited participants and collected clinical data. Ryoji Goto, Harutsugu Tatebe, Ayami Okuzumi, Gen Matsumoto, Kenji Tagai, Sayo Matsuura, Wataru Satake, Tatsushi Toda, Yuko Kataoka, Hironobu Endo, Makoto Higuchi, and Takahiko Tokuda interpreted the data. Ryoji Goto drafted the initial manuscript. All authors contributed to revision and editing of the manuscript.

## Acknowledgments

We would like to thank all the subjects for participating in this study. We would like to express our sincere gratitude to Ms. Miyuki Yoshiya for her assistance with the immunoblotting experiments, and to Ms. Kumiko Yamaguchi for her support in the procurement of reagents.

## Notes

### Author Declarations

The Institutional Review Board of the National Institutes for Quantum Science and Technology gave ethical approval for this work.

