## Supplementary Materials for "Plasma acetylated α-synuclein as a novel quantitative biomarker for Parkinson’s disease"

**eMethods**

### Generation of a monoclonal antibody specific to N-terminally acetylated α-synuclein

#### 1. Animals

All animal procedures were approved by the Committee for Animal Experiments at Japan SLC, Inc. (Shizuoka, Japan) and by Cell Engineering Corporation (Osaka, Japan). A Japanese white rabbit (JW/CSK) was supplied, immunized, and euthanized for lymph node collection by Japan SLC, Inc.

#### 2. Production of rabbit monoclonal antibodies against acetylated α-synuclein

A synthetic N-terminally acetylated α-synuclein peptide was conjugated to keyhole limpet hemocyanin (KLH) and used as the immunogen. The rabbit was immunized subcutaneously with the KLH–peptide conjugate emulsified in Freund's complete adjuvant, followed by a booster injection without adjuvant after two weeks. Seven days after the booster, the rabbit was euthanized and draining lymph nodes were collected. Lymphocytes were isolated, suspended in cryopreservation medium, and stored at −80˚C.

Cryopreserved lymphocytes were applied to microchamber array chips pre-coated with peptide–BSA conjugates of the acetylated peptide for antigen-specific single B-cell screening. Secreted antibodies were detected by on-chip fluorescence immunoassay, and antigen-reactive B cells were individually isolated using an automated micromanipulator (AS ONE, Osaka, Japan). Total RNA was extracted, and VH and VL genes were amplified by RT-PCR. The paired VH and VL sequences were subcloned into expression vectors containing rabbit IgG constant-region genes, and the resulting constructs were co-transfected into HEK293 cells to produce recombinant antibodies. Culture supernatants were screened by enzyme-linked immunosorbent assay (ELISA), and clones exhibiting selective reactivity toward acetylated α-synuclein were selected. Selected antibody constructs were transiently expressed in suspension-adapted CHO cells, and antibodies were purified using protein A affinity chromatography.

#### 3. ELISA

ELISA was performed to evaluate antibody reactivity and selectivity for acetylation. Plates were coated with peptide–BSA conjugates of either the acetylated peptide or the non-acetylated peptide. After blocking, culture supernatants were added, and bound antibodies were detected using HRP-conjugated anti-rabbit IgG followed by TMB substrate development. Clones showing preferential binding to the acetylated peptide were selected for further analysis.

### **Immunoblotting analysis**

Immunoblotting was performed to verify the specificity of antibodies recognizing the N-terminal acetylation epitope of human α-synuclein. Serial dilutions of recombinant N-terminally acetylated human α-synuclein (StressMarq, #SPR-331) and protein standards (Bio-Rad, #1610374) were resolved by SDS-PAGE on a precast gradient gel (Nacalai Tesque) and transferred to a nitrocellulose membrane (Bio-Rad, #1620115). After blocking with TBS containing 5% skim milk and 0.05% Tween-20, membranes were incubated with the primary antibody (rabbit monoclonal antibody against N-terminally acetylated α-synuclein, diluted 1:1,000; as described above), washed to remove unbound antibody, and probed with an HRP-conjugated secondary antibody. Chemiluminescence was generated using Western Lightning Plus ECL (PerkinElmer, #ORT2755) and imaged with an Amersham Imager 680QC (Cytiva).

**Procedures for immunoassay method validation**

Immunoassay method validation was conducted according to the guiding principles reported previously [1,2].

1. **Determination of the limit of detection (LOD) and lower limit of quantification (LLOQ)**

Eight aliquots of blank samples were prepared, and the "background" signals of our novel plasma acetylated α-synuclein immunoassay on the Simoa HD-X analyzer (Quanterix) were measured. In Simoa, the measured signals were quantified as AEB values. The LOD was determined as the interpolated acetylated α-synuclein concentration corresponding to the mean plus 2.5 standard deviations (SDs) of the AEB values for blank samples. The LLOQ was determined as the interpolated acetylated α-synuclein concentration corresponding to the mean plus 10 SDs of the AEB values for blank samples.

2. **Intra-assay precision**

Twenty-four samples with different concentrations of calibrators were prepared for the analysis of intra-assay precision, and the concentrations were measured in one experiment. The intra-assay precision was determined by calculating the within-run coefficient of variation (CV) of the measured concentrations of these calibrators.

### 3. Inter-assay precision

The inter-assay precision was determined by calculating the coefficient of variation (CV) of the concentrations derived from three plasma samples of different concentrations and from two calibrators. The acetylated α-synuclein levels in these samples were measured four times on different days. The inter-assay precision was determined by calculating the CV of the concentrations across runs for those samples.

### 4. Spike recovery tests, parallelism, and dilution linearity experiments

For the spike recovery tests, four aliquots of each of two plasma samples with different acetylated α-synuclein concentrations were prepared and spiked with 0, 250, 500, and 1000 pg/mL of calibrator peptide in 160 μL of solution (containing plasma, sample buffer, and spiked peptide). These eight (4 × 2) aliquots were analyzed in duplicate in the same run. Two spike recovery curves were generated, ranging from the non-spiked solution to the 1,000 pg/mL-spiked solution, to evaluate parallelism. Recovery rates (% recovery) were calculated by correcting the measured concentration for the endogenous calibrator peptide concentration.

To assess dilution linearity, the high-concentration spiked sample prepared in the spike recovery tests described above was used, and twofold serial dilutions (× 2, × 4, and × 8) of a spiked sample were generated using the sample diluent until the theoretical concentration reached the LLOQ. Serially diluted samples were analyzed in duplicate.

**eResults**

### Immunoblotting analysis of the reactivity of our novel antibody specific to N-terminally acetylated α-synuclein

Immunoblotting analysis using our novel anti–acetylated α-synuclein antibody showed a distinct band at the expected molecular weight (around 18 kDa) detected in Lane 1 (arrow, 1,000 ng/lane), whereas no visible bands were observed in the lower-concentration lanes (Lanes 2, 3, and 4: 100, 10, and 1 ng/lane, respectively), as shown in eFigure 1. This pattern reflects the lower analytical sensitivity of immunoblotting compared with our immunoassay (LOD = 50 pg/mL). Notably, the loaded protein amounts represent total α-synuclein, of which only less than 10% is acetylated; therefore, the effective amount of acetylated α-synuclein in each lane falls below the level required for visual detection. These findings are thus consistent with the expected detection characteristics and support selective recognition of N-terminally acetylated α-synuclein.

### Standard curve for the novel acetylated α-synuclein immunoassay and assay method validation

A representative standard curve for our novel acetylated α-synuclein immunoassay is shown in eFigure 2. Using our novel assay, acetylated α-synuclein could be detected with high sensitivity and with a wide dynamic range of approximately 500–10,000 pg/mL.

LOD of the assay was 59.7 pg/mL, calculated using the calibrator peptide (synthesized peptide containing the acetylation site in the N-terminal region and mid-portion of α-synuclein; molecular weight: 3313.3 Da). Based on molecular weight normalization to full-length acetylated α-synuclein (molecular weight: 14,502 Da), the

LOD was equivalent to 261 pg/mL for acetylated α-synuclein. The lower limit of quantification (LLOQ) of the assay was 464 pg/mL for the calibrator peptide, or 2,032 pg/mL for acetylated α-synuclein. Intra-assay precision was robust, with CVs between 3.4% and 7.8% (eTable 1). The %CV of the AEBs at different concentrations was between 2.8% and 9.3% (eTable 2), indicating that our novel acetylated α-synuclein assay showed good inter-assay precision (< 10%). In the spike recovery and parallelism experiments, the recovery rate (%Recovery) of each sample was 94.6%–139.2% (eTable 3). The parallelism of two spike recovery curves ranging from the non-spiked solution to the 1,000 pg/mL-spiked solution is also shown in eFigure 3, which demonstrated that plasma samples spiked with 0, 250, 500, and 1,000 pg/mL of the calibrator peptide provided reliable results and parallelism. The dilution linearity experiments demonstrated that a sample with a spiked concentration could be diluted to a concentration within the working range and still give a reliable quantification. eFigure 4 shows the results of those experiments, providing reliable quantification after dilution within the standard curve range (eTable 4).

**eTable 1. Intra-assay precision (n = 24)**

| | Sample 1: 500 pg/mL (calibrator peptide) | Sample 2: 1000 pg/mL (calibrator peptide) | Sample 3: 2000 pg/mL (calibrator peptide) | Sample 4: 4000 pg/mL (calibrator peptide) |
|---|---|---|---|---|
| Mean AEB | 0.159 | 0.325 | 0.834 | 3.159 |
| SD | 0.00533 | 0.0133 | 0.0649 | 0.109 |
| CV (%) | 3.4 | 4.1 | 7.8 | 3.4 |

SD, standard deviation; CV, coefficient of variation.

**eTable 2. Inter-assay precision (n = 20)**

| | Sample 1 (plasma) | Sample 2 (plasma) | Sample 3 (plasma) | Sample 4 (calibrator peptide) | Sample 5 (calibrator peptide) |
|---|---|---|---|---|---|
| Mean AEB | 1.45 | 0.208 | 0.186 | 13.182 | 4.340 |
| SD | 0.115 | 0.013 | 0.005 | 1.219 | 0.323 |
| CV (%) | 7.9 | 6.2 | 2.8 | 9.3 | 7.4 |

SD, standard deviation; CV, coefficient of variation.

**eTable 3. Recovery rates (% recovery) for each plasma sample in spike recovery tests**

| | Treatment | Measured concentration (pg/mL) | Concentration CV (%) | Theoretical concentration of the spiked sample (pg/mL) | Recovery rate (%) |
|---|---|---|---|---|---|
| Sample 1 | Neat (0 pg/mL spike) | 881.5 | 1.1 | | |
| | 250 pg/mL spike | 1130 | 3.0 | 1131 | 99.7 |
| | 500 pg/mL spike | 1392 | 3.8 | 1381 | 102.2 |
| | 1000 pg/mL spike | 1828 | 1.3 | 1881 | 94.6 |
| Sample 2 | Neat (0 pg/mL spike) | 473.7 | 20.5 | | |
| | 250 pg/mL spike | 821.7 | 0.0 | 723.7 | 139.2 |
| | 500 pg/mL spike | 1030 | 6.9 | 973.7 | 111.2 |
| | 1000 pg/mL spike | 1611 | 2.6 | 1473.7 | 113.8 |

CV, coefficient of variation.

**eTable 4. Dilution linearity of a spiked sample**

| | Fold dilution | Mean measured concentration (pg/mL) | Concentration CV (%) | Dilution corrected concentration (pg/mL) | Recovery rate (%) |
|---|---|---|---|---|---|
| Sample | 2 | 2795.3 | 1.1 | 2795.3 | 100.0 |
| | 4 | 1611.2 | 2.6 | 3222.5 | 115.3 |
| | 8 | 1051.8 | 0.9 | 4207.1 | 150.5 |

CV, coefficient of variation.

**eTable 5. Correlations between plasma acetylated α-synuclein levels and clinical variables in patients with PD.**

| | age | disease duration | MDS-UPDRS part3 | Yahr | MIBG cardiac scintigraphy [2] early acquisition | MIBG cardiac scintigraphy [2] late acquisition | MIBG cardiac scintigraphy [2] washout ratio | DAT SPECT SBR | MMSE | MoCA-J | FAB | Open Essence [1] |
|---|---|---|---|---|---|---|---|---|---|---|---|---|
| *r*= | 0.04 | 0.16 | 0.12 | 0.13 | −0.14 | −0.20 | −0.16 | 0.01 | −0.10 | 0.12 | 0.11 | −0.23 |
| *p*= | 0.77 | 0.26 | 0.39 | 0.36 | 0.34 | 0.19 | 0.29 | 0.97 | 0.48 | 0.41 | 0.45 | 0.10 |

The items were analyzed in [1] 51 and [2] 46 patients.

MMSE, Mini-Mental State Examination; MoCA-J, Japanese version of Montreal Cognitive Assessment; FAB, Frontal Assessment Battery; MDS-UPDRS, Movement Disorder Society-Unified Parkinson's Disease Rating Scale; DAT SPECT, dopamine transporter single-photon emission computed tomography; SBR, specific binding ratio.

**eTable 6. Correlations between CSF acetylated α-synuclein levels and clinical variables in patients with PD.**

| | | age | disease duration | MDS-UPDRS part3 | Yahr |
|---|---|---|---|---|---|
| PD (n = 51) | n = | 51 | 51 | 51 | 51 |
| | *r* = | –0.2547 | 0.008812 | –0.1007 | 0.01459 |
| | *p* = | 0.0713 | 0.9511 | 0.4818 | 0.9191 |

MDS-UPDRS, Movement Disorder Society-Unified Parkinson's Disease Rating Scale.

**eTable 7. Correlations between CSF acetylated α-synuclein levels and clinical variables in patients with MSA.**

| | | age | disease duration | MDS-UPDRS part3 | Yahr | ICARS | MMSE |
|---|---|---|---|---|---|---|---|
| MSA total (n = 25) | n = | 25 | 25 | 10 | 25 | 10 | 19 |
| | *r* = | 0.396 | 0.2815 | 0.1879 | 0.1554 | 0.07879 | **–0.601** |
| | *p* = | 0.0501 | 0.1728 | 0.6073 | 0.4583 | 0.8382 | **0.0065** |
| MSA-P only (n = 8) | n = | 8 | 8 | 7 | 8 | | 8 |
| | *r* = | 0.6454 | 0.5608 | 0.2857 | 0.0894 | | –0.4392 |
| | *p* = | 0.0839 | 0.1482 | 0.556 | 0.8452 | | 0.2875 |
| MSA-C only (n = 17) | n = | 17 | 17 | 3 | 17 | 10 | 11 |
| | *r* = | 0.3665 | 0.2433 | –1.0000 | 0.2205 | 0.07879 | –0.5987 |
| | *p* = | 0.1479 | 0.3436 | 0.3333 | 0.4324 | 0.8382 | 0.0559 |

MDS-UPDRS, Movement Disorder Society-Unified Parkinson’s Disease Rating Scale; ICARS, International cooperative ataxia rating scale; MMSE, Mini-Mental State Examination.

**eFigure 1. Validation of monoclonal antibody specificity for the acetylated N-terminus of human α-synuclein.**

Serial dilutions of recombinant N-terminally acetylated human α-synuclein together with protein standards were separated by SDS-PAGE and immunoblotted using a rabbit monoclonal anti–acetylated α-synuclein antibody (clone 8; 1:1,000). Each lane contained the indicated amount of recombinant protein (Lane 1: 1,000 ng; Lane 2: 100 ng; Lane 3: 10 ng; Lane 4: 1 ng). A distinct immunoreactive band at approximately 18 kDa (arrow) corresponds to the expected molecular weight of acetylated α-synuclein.

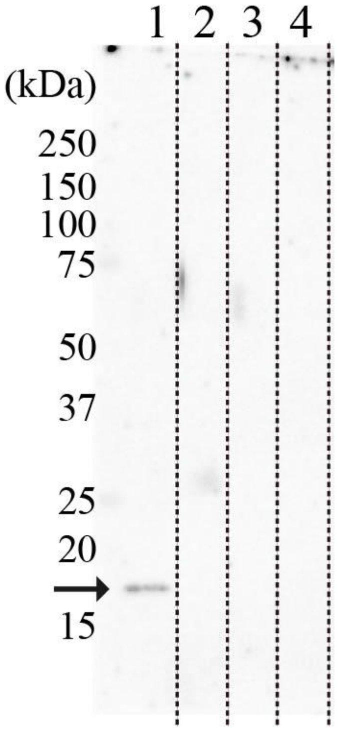

**eFigure 2. Representative standard curve for the novel acetylated α-synuclein immunoassay.**

A representative standard curve for our novel acetylated α-synuclein immunoassay is shown, displaying the AEB signal on the vertical axis and the known concentrations of the calibrator on the horizontal axis. The curve was generated by fitting the data with a four-parameter logistic model. The goodness of fit ($R^2$) of the standard curve was 0.9995. The assay is based on the ultrasensitive digital array technology of the Simoa system (Quanterix). Data represent the mean ± SD of duplicate readings.

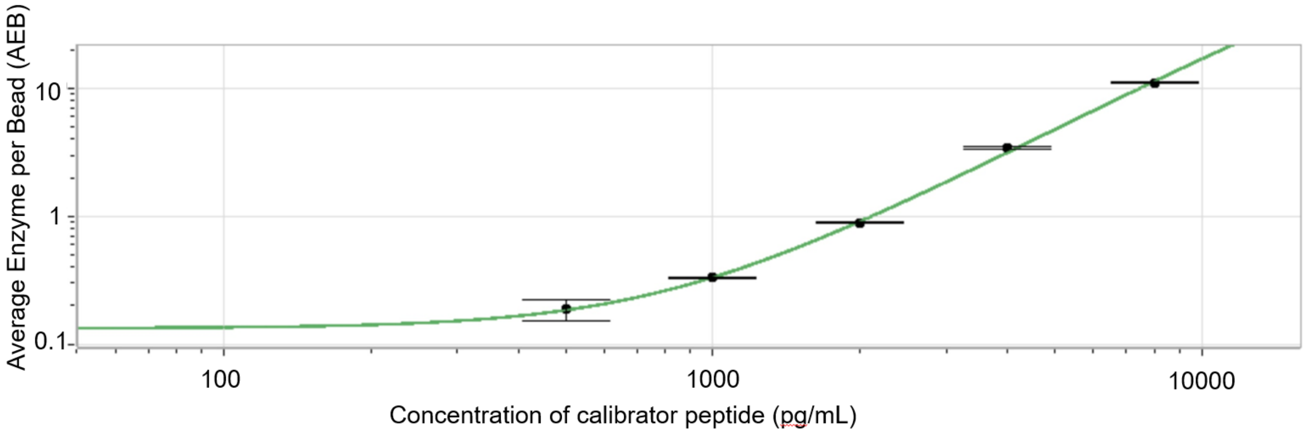

**eFigure 3. Recovery rates (% recovery) for each plasma sample in the spike recovery tests**

Four aliquots from each of two plasma samples with different acetylated α-synuclein concentrations were spiked with 0, 250, 500, and 1,000 pg/mL of the calibrator peptide in 160 μL of solution. These eight (4 × 2) aliquots were analyzed in duplicate. Two spike recovery curves ranging from the non-spiked solution to the 1,000 pg/mL-spiked solution were generated to evaluate parallelism. Recovery rates (% recovery) were calculated by correcting the measured concentration for the endogenous calibrator peptide concentration.

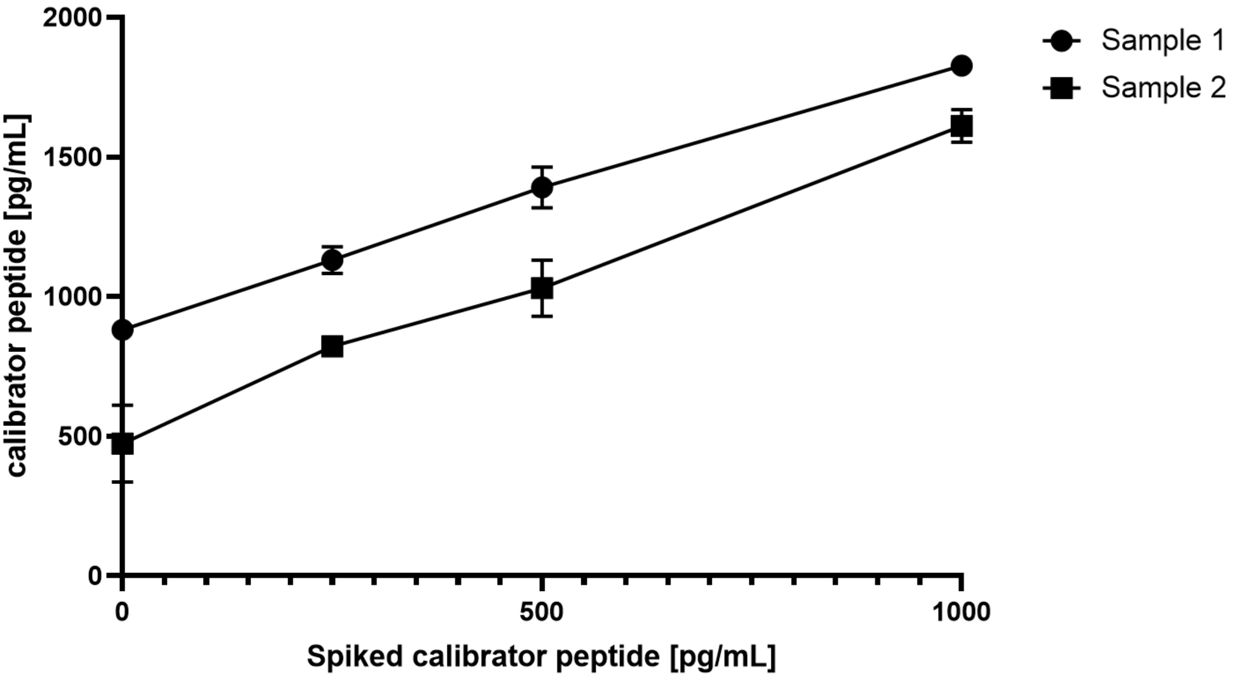

**eFigure 4. Dilution linearity of a spiked sample**

The high-concentration spiked sample prepared in the spike recovery tests was used, and twofold serial dilutions (×2, ×4, and ×8) of the spiked sample were generated using the sample diluent until the theoretical concentration reached the lower limit of quantification (LLOQ). Serially diluted samples were analyzed in duplicate.

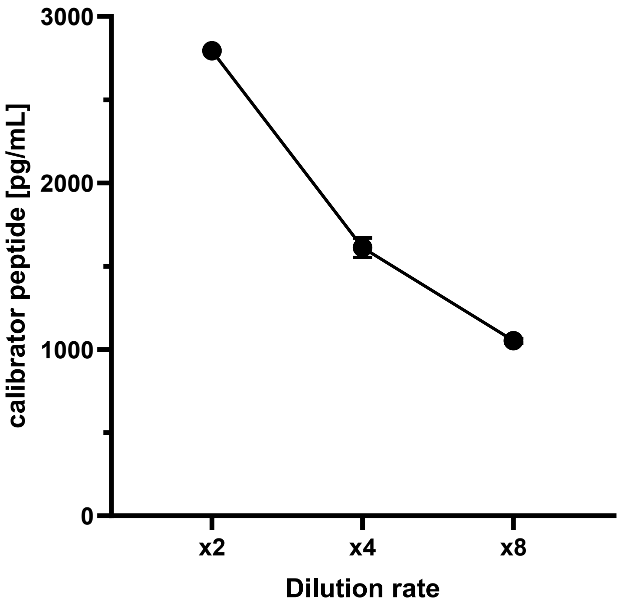

**eFigure 5. CSF acetylated α-synuclein levels in patients with MSA.**

No significant difference in acetylated α-synuclein levels was observed between MSA-P and MSA-C groups ($p$ = 0.3440).

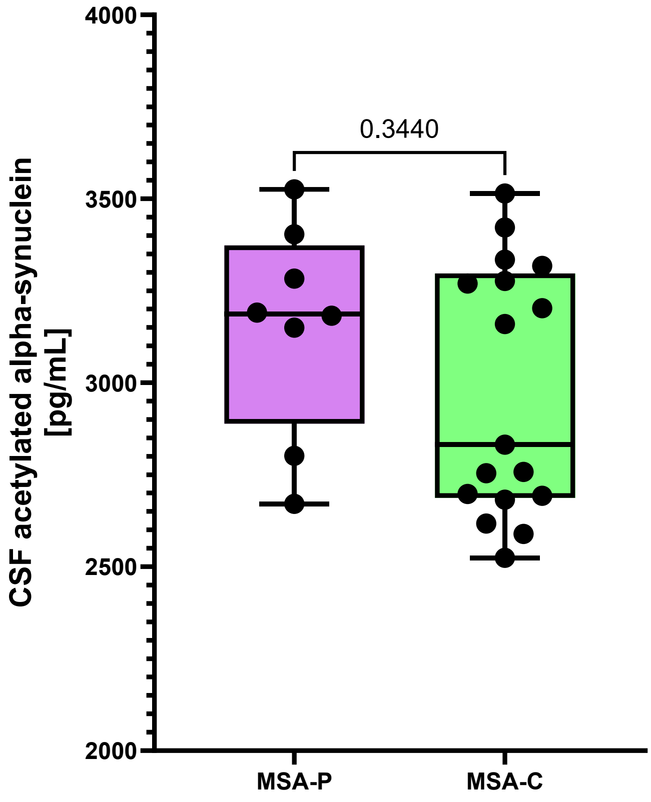

**eFigure 6. Correlation between CSF acetylated α-synuclein levels and Mini-Mental State Examination (MMSE) scores in MSA.**

In the analysis of all patients with MSA, CSF acetylated α-synuclein levels correlated with MMSE scores ($r = -0.601$, $p = 0.0065$).

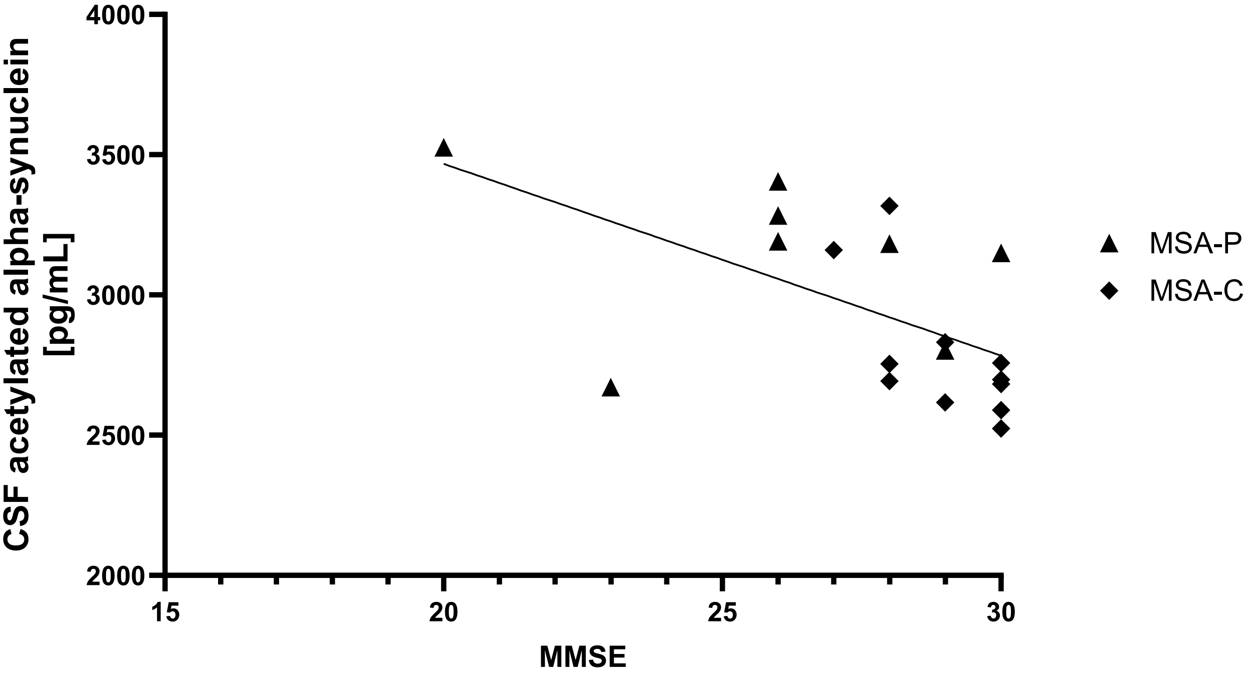

**eFigure 7. CSF and plasma acetylated α-synuclein levels in patients with PD and MSA.**

Acetylated α-synuclein levels in CSF and non-hemolyzed plasma did not correlate in patients with PD ($r = -0.12$, $p = 0.65$) or in those with MSA ($r = -0.10$, $p = 0.86$).

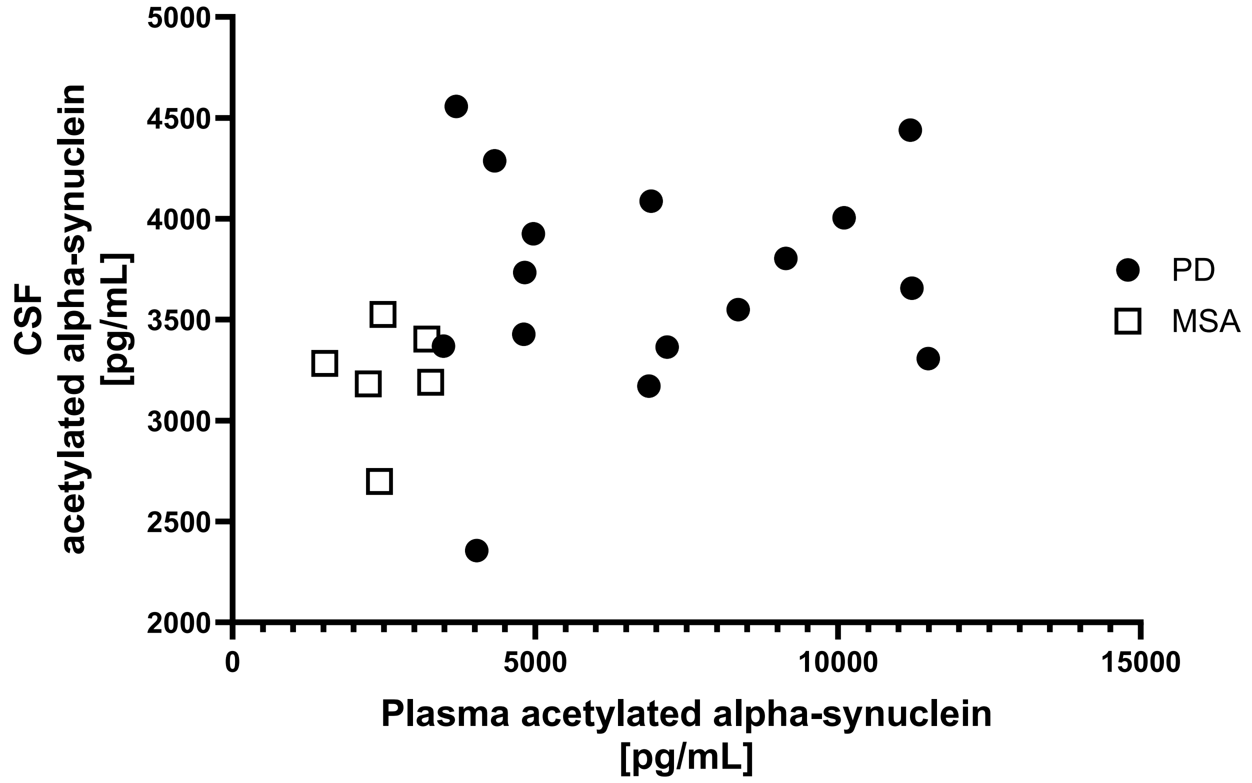
